# Negative-Binomial and Quasi-Poisson regressions between COVID-19, mobility and environment in São Paulo, Brazil

**DOI:** 10.1101/2021.02.08.21250113

**Authors:** Sergio Ibarra-Espinosa, Edmilson Dias de Freitas, Karl Ropkins, Francesca Dominici, Amanda Rehbein

**Author notes:** **Corresponding author** Sergio Ibarra-Espinosa, Departamento de Ciências Atmosféricas, Instituto de Astronomia, Geofísica e Ciências Atmosféricas, Universidade de São Paulo, Brazil.

## Abstract

Brazil, the country most impacted by the coronavirus disease 2019 (COVID-19) on the southern hemisphere, use intensive care admissions per day, mobility and other indices to monitor quarantines and prevent the transmissions of SARS-CoV-2. In this study we quantified the associations between residential mobility index (RMI), air pollution, meteorology, and daily cases and deaths of COVID-19 in São Paulo, Brazil. We applied a semiparametric generalized additive model (GAM) to estimate: 1) the association between RMI and COVID-19, accounting for ambient particulate matter (PM_2.5_), ozone (O_3_), relative humidity, temperature and delayed exposure between 3-21 days, and 2) the association between COVID-19 and exposure to for ambient particulate matter (PM_2.5_), ozone (O_3_), accounting for relative humidity, temperature and mobility. We found that an RMI of 45.28% results in 1,212 cases (95% CI: 1,189 to 1,235) and 44 deaths (95% CI: 40 to 47). Increasing the isolation from 45.28% to 50% would avoid 438 cases and 21 deaths. Also, we found that an increment of 10 μg·m^-3^ of PM_2.5_ results in a risk of 1.140 (95% CI: 1.021 to 1.274) for cases and 1.086 (95% CI: 1.008 to 1.170) for deaths, while O_3_ produces a relative risk of 1.075 (95% CI: 1.006 to 1.150) for cases and 1.063 (95% CI: 1.006 to 1.124) for deaths, respectively. We compared our results with observations and literature review, finding well agreement. Policymakers can use such mobility indices as tools to control social distance activities. Spatial distancing is an important factor to control COVID-19. Small increments of air pollution result in an increased number of COVID-19 cases and deaths.

## 1. Introduction

The world has been facing an unprecedent critical health crisis due to COVID-19 pandemic caused by the severe acute respiratory syndrome coronavirus 2 (SARS-CoV-2). According to the Johns Hopkins University (JHU) Center for Systems Science and Engineering (CSSE), on February 20^th^, 2021, there were 110,810,601 confirmed cases of COVID-19 worldwide, and 2,454,047 deaths (Dong et al. 2020). Many countries implemented social isolation and quarantine strategies, while internet companies released anonymized aggregated location data to provide information on the effectiveness of quarantine and isolation (Google 2021). This new disease demonstrated that developed countries such as Belgium, Italy, and Spain were unprepared, which resulted in infection fatality rates (deaths/cases) of 15.4%, 13.5%, and 10.2%, respectively. Furthermore, new mutations of SARS-CoV-2 would present higher transmissibility and current vaccines might not offer protection against it (Gupta et al. 2021; Kupferschmidt 2021). Therefore, it is urgent and crucial to conduct more research to better understand the relationships and associations between SARS-CoV-2 transmissibility and environmental factors.

COVID-19 spread very quickly across Latin America (Bolaño-Ortiz et al. 2020; Tello-Leal and Macías-Hernández 2020). As of January 4, 2021, Brazil, with an area of 8.5 million km^2^ and a population of 208 million, is the third country with more cases (7,733,746) and deaths (196,018) (Dong et al. 2020). The city of São Paulo, the most populated city in Brazil with 11.8 million inhabitants (IBGE 2020), presents the highest number of COVID-19 cases (404,025) and deaths (15,725, https://covid.saude.gov.br/). In Brazil, only symptomatic cases are tested, hence, the real number of SARS-CoV-2 infections could be much higher. A recent study shows that COVID-19 death notification in Brazil is underreported (Alves et al. 2020). On March 24^th^, 2020 (GESP 2020), São Paulo’s government recommended social distancing for suspected cases and introduced a local quarantine to reduce virus transmission. Brazil adopted containment measures such as close contact and limited mobility as protective measures, however “quarantine” was the official term used by São Paulo state government. Consequently, air pollution levels dropped during the quarantine. For instance, the concentrations of carbon monoxide (CO), nitrogen dioxide (NO_2_), PM_10,_ and PM_2.5_ were reduced by -35.70%, -29.56%, -17.80%, and -25.02%, respectively, while O_3_ increased by 53.25% between 21:00-03:00 Local Time (LT) (Dantas et al. 2020; Freitas et al. 2020; Nakada and Urban 2020). Other studies showed reductions in China (30% of NO_2_ and 25% of CO_2_) (Dutheil et al. 2020). However, there are indications that COVID-19 and air pollution interactions may be more complex. Some early reports suggest that longer-term exposure to air pollution increases susceptibility and severity upon infection (Tosepu et al. 2020; Wu et al. 2020). Similarly, meteorological interactions are likely to be important, for example, some studies have suggested that high humidity and temperature would reduce virus transmission (Wang et al. 2020a, 2020b). Furthermore, the biological inactivation rate of SARS-CoV-2 is sensitive to humidity, with minimum half-life at 65% and higher at 40% and 85%, and its decays at 27 °C is 10 times faster than at 10 °C (Morris et al. 2020). However, estimations for the Amazonian city of Manaus, which have a climatology^1^ temperature 27 °C of and relative humidity 80.2% for July, show that between 44% and 66% of the population was infected with SARS-CoV-2 by July (Buss et al. 2020). Then, more research is needed to better understand the interactions between SARS-COV-2 and environmental factors.

Some studies have evaluated the effect of mobility on COVID-19, mainly in China. Kraemer et al., (2020) found correlation of 0.94 between real-time mobility data and COVID-19 cases from Wuhan, China, confirming the exportation of cases from Wuhan to other provinces and the effectiveness of the sanitaire cordon. Tian et al (2020) found that Wuhan shutdown delayed the appearance of COVID-19 in 2.9 days. Zhu et al (2020b) found that increasing the human mobility increased 6.45% daily confirmed cases. In the US, Badr et al (2020) associated county-level origin-destination matrices and COVID-19 cases finding that counties with more mobility presented higher number of cases. In a related correspondence, Gatalo et al (2020) found that the absence of a strong correlation between growth of cases and mobility may be related to other factors, such as wearing mask, keeping distance in encounters and also, the existence of superspreading events.

Our study investigates the associations between RMI and COVID-19, and air pollution and COVID-19 on a particular day, having accounted for environmental and meteorological factors. We analyzed the effects of mobility, air pollution, and meteorology on the daily COVID-19 cases and deaths in the city of São Paulo. We applied semiparametric generalized additive models to study the effect of each predictor by isolating confounding factors (Peng and Dominici 2008). To the best of our knowledge, few studies have associated mobility air pollution, meteorological factors, and COVID-19 simultaneously especially in Latin America.

## 2. Material and Methods

The Brazilian Ministry of Health reports the official daily time-series of cases and mortality associated with COVID-19 at https://covid.saude.gov.br/ updated once a day around 19:00 Brazilian official time (−3 GMT). This information is gathered from the 26 states and the federal district health secretaries and provided at the national, state, and municipality levels. The availability of data using the web site was interrupted on June 07^th^, 2020, and returned on June 08^th^, 2020 after judicial demand (G1 2020). When returned, it included the data obtained when the web site was offline. To check the trustworthiness of that data, we compared it with the dataset from BrasilIO (https://brasil.io/covid19/). The BrasilIO is an independent organization made of voluntaries that also gathers COVID-19 data from state health secretaries. Both datasets are very similar, in effect, we applied the Wilcoxon test (R Core Team 2021) for cases and deaths getting p-values of 0.7798 and 0.922, meaning that there are no significant differences between the two datasets. Therefore, we used the official data from the Brazilian Ministry of Health. The day July 29^th^ the number of cases reported included July 28^th^; therefore, we distributed the number of cases and deaths equally in both days. Figure 1 shows the daily cases and deaths from COVID-19 in São Paulo State between March 27^th^, 2020 and January 03^rd^, 2021. The daily mean cases are 1432 and deaths is 55, with a maximum of 7063 on August 13^rd^, 2020 and 179 on June 23^rd^, 2020, respectively. Furthermore, the variances are 1511183 for cases and 1889.89 for deaths, indicating overdispersion in the data. In effect, the dispersion parameter for cases is 1035.86 and for deaths 33.84 (both p-value<0.05) (Kleiber and Zeileis 2008).

**Figure 1.**
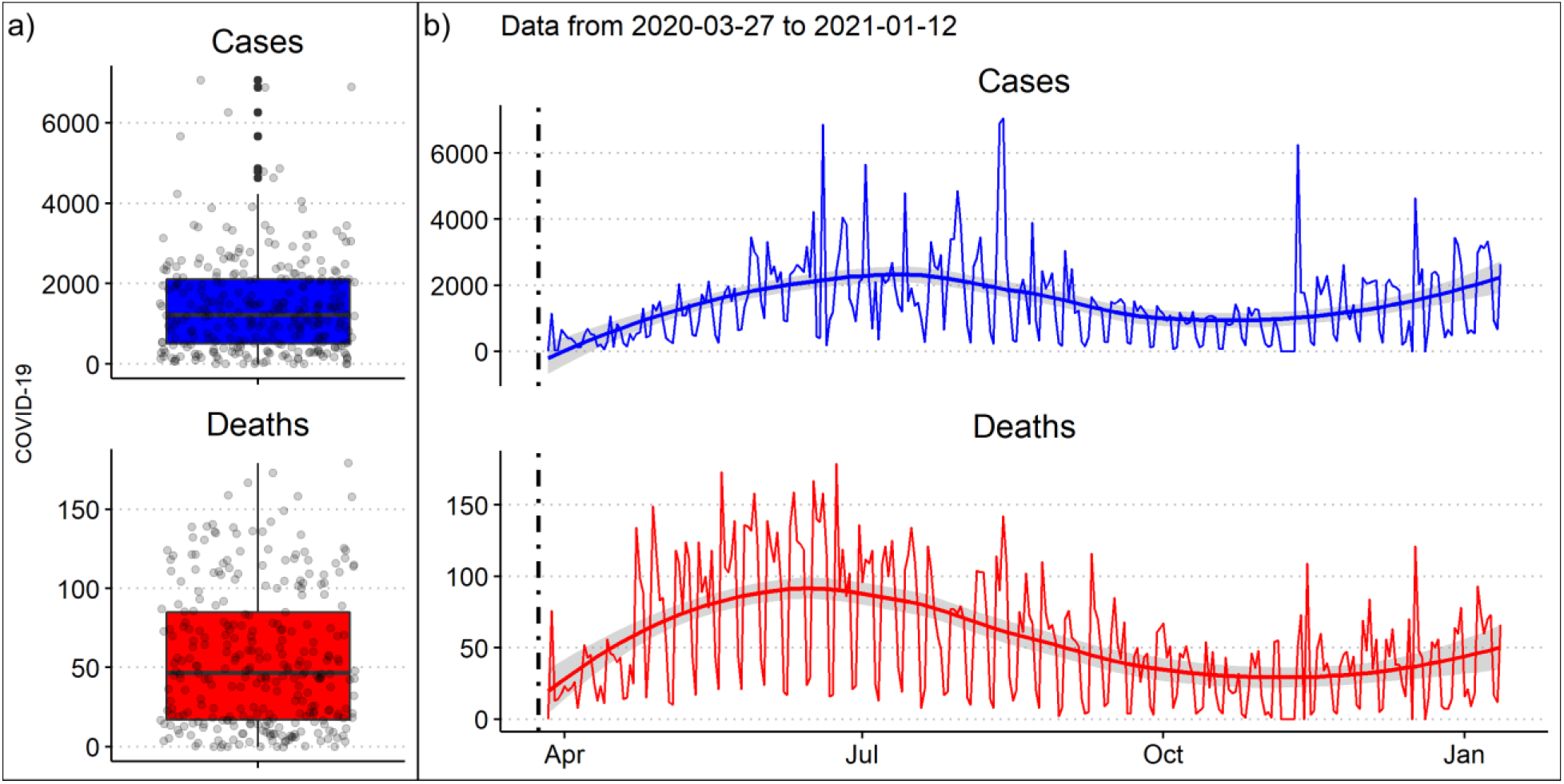
Daily cases and deaths of COVID-19 in (a) boxplots and (b) time series in São Paulo, Brazil, between March 27^th^, 2020 and January 12^nd^, 2021 (Saúde 2021). The smooth lines on panel b) are the LOESS regression made with ggplot2 and R (R Core Team 2021; Wickham 2016).

We obtained mobility datasets from Google mobility trends (Google, 2020) and the Intelligent Monitoring System for the city of São Paulo (SIMI-SP). The Google data is based on the use of smart devices such as cellphones, vehicle trackers, and other GPS enabled systems^2^. The data reported by Google consists in mobility trends related to places as percentage change from baseline, with baseline as the median value, for each day of the week, during the 5-week period January 3^rd^ and February 6^th^, 2020. We selected the mobility trend “residential”, because it better represent the situation of *spatial isolation* (Hereafter Residential Mobility Index, RMI), then, higher values of RMI means higher isolation. The SIMI-SP data is collected by network companies which receive cellphone triangulating signals with the nearest cellphone communication tower^3^. This means that SIMI-SP does not need internet to collect mobility information. Both indices are shown on Figure 2, meaning that higher values represent more stay-at-home and while lower values pre-quarantine conditions. RMI is an indicator that represent exposure in two ways, first, staying out of home increases the chances of getting infections with SARS-CoV-2 present in aerosols (Morawska and Milton 2020) and second, increases the exposure to air pollution with deleterious effect on human health which could pose a synergetic effect. The trend on both RMI shows steady decline ahead of the start of quarantine on March 24^th^, 2020 (black vertical line) (GESP 2020). The median RMI for Google data 13.51% and for RMI SIMI-SP is 44.75% for the whole period. The median RMI for the pre-quarantine period (first 15 days of March 2020), are -1% for Google and 31.30% for SIMI-SP and after the quarantine 14% for Google and 45.48% for SIMI-SP.

**Figure 2.**
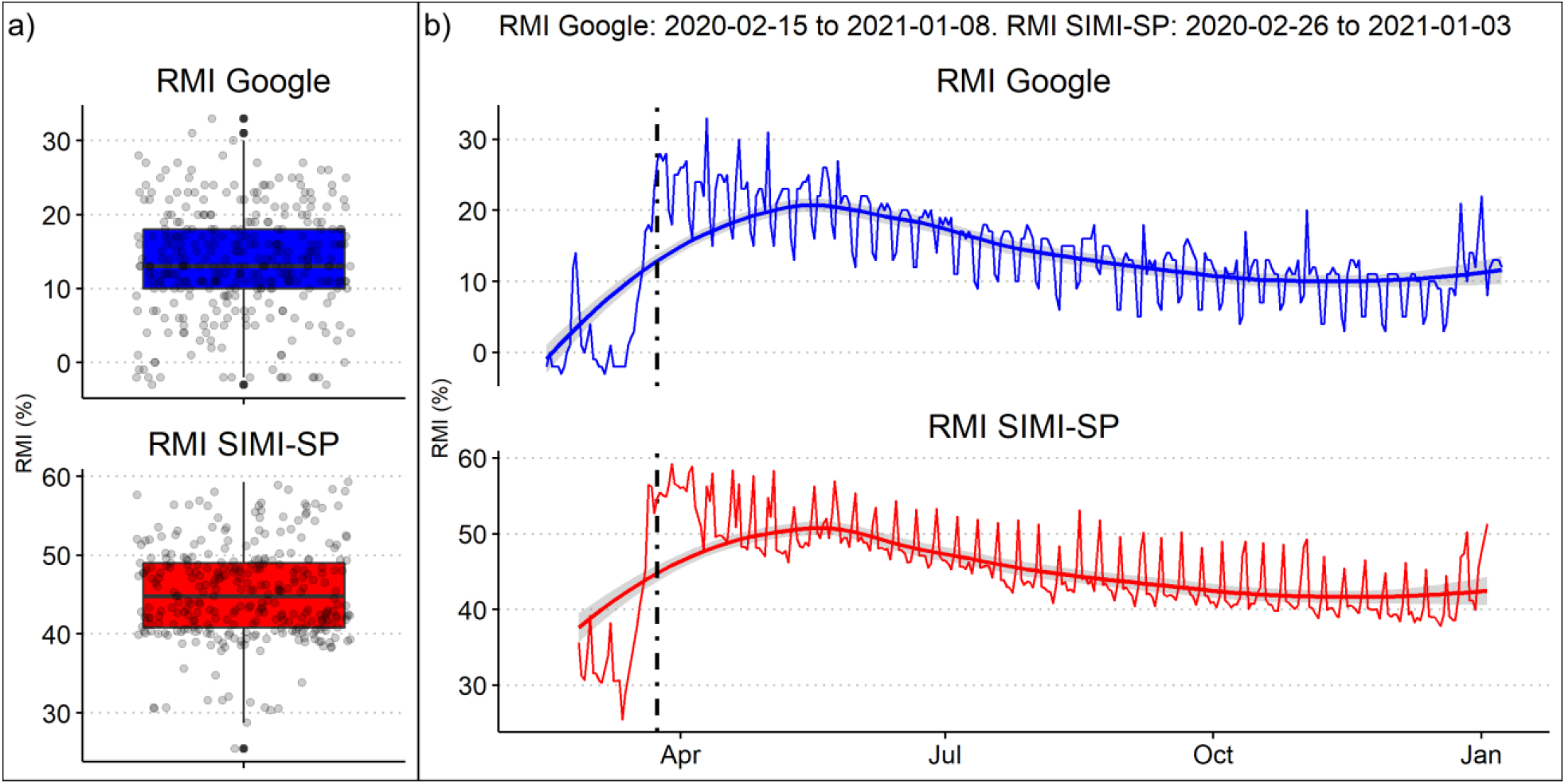
RMI values in São Paulo, Brazil, in a) boxplots and b) times series between January 15^th^ and December 29^th^, 2020 for RMI Google (Google 2021) and between February 26, 2020 and January 3, 2021. Black vertical line shows when started the quarantine in São Paulo on March, 24. The smooth lines on panel b) are the LOESS regression made with ggplot2 and R (R Core Team 2021; Wickham 2016).

Air pollution and meteorology hourly surface measurements were obtained from the air quality system (QUALAR) operated by the Environmental Agency of Sao Paulo State (CETESB, 2020). QUALAR archives air quality real-time data using several stations spread in São Paulo. Hourly averages of O_3_ (ug/m^3^), PM_2.5_ (ug/m^3^), relative humidity (%) and temperature (°C) are shown in Figure 3, between January 01^st^, 2019 and January 4^th^, 2021. Locally estimated scatterplot smoothing (or local regression LOESS) show that that the O_3_ incremented after the quarantine and remained higher than 2019 during most of 2020. The concentrations of PM2.5 during 2020 were lower than 2019, related to the decrease in human activity and also, as reported by other studies (Bolaño-Ortiz et al. 2020; Debone et al. 2020; Nakada and Urban 2020), and just in September concentration increased. The quarantine beginning coincided with the dry season beginning at São Paulo, presenting a decline in temperature compared to the early months of 2020. The most significant feature of the São Paulo dry season (April to November), the wet season counterpart (Vera et al. 2006), is the non-significative precipitation amount, caused by weak isolated events, and long periods with no precipitation occurring in between (Rehbein et al. 2018), and the very low relative humidity during the daytime (climate reports from the Climate Group of Studies - GrEC/USP, http://www.grec.iag.usp.br). Also, a general decreasing in temperature occurs according to the austral winter and presenting drops in temperature, associated to the synoptic systems crossing São Paulo and generally are not able to organize convection (GrEC/USP, http://www.grec.iag.usp.br) or even sea breezes that eventually reaches the interior of São Paulo (Freitas et al. 2007). In 2020, for instance, from April 3^rd^ -May 3^rd^, 2020 there was very few (2 mm at São Paulo-SESC Interlagos station, in the south of São Paulo) or no precipitation (at the São Paulo-Mirante de Santana station, in the north of São Paulo) according to the official meteorological stations (www.inmet.gov.br), while synoptic systems (such as cold fronts) were observed (CPTEC/INPE, https://www.cptec.inpe.br; GrEC/USP, http://www.grec.iag.usp.br). The first semester of 2020 was drier and colder than 2019, but during the second semester, relative humidity and temperature remained similar.

**Figure 3.**
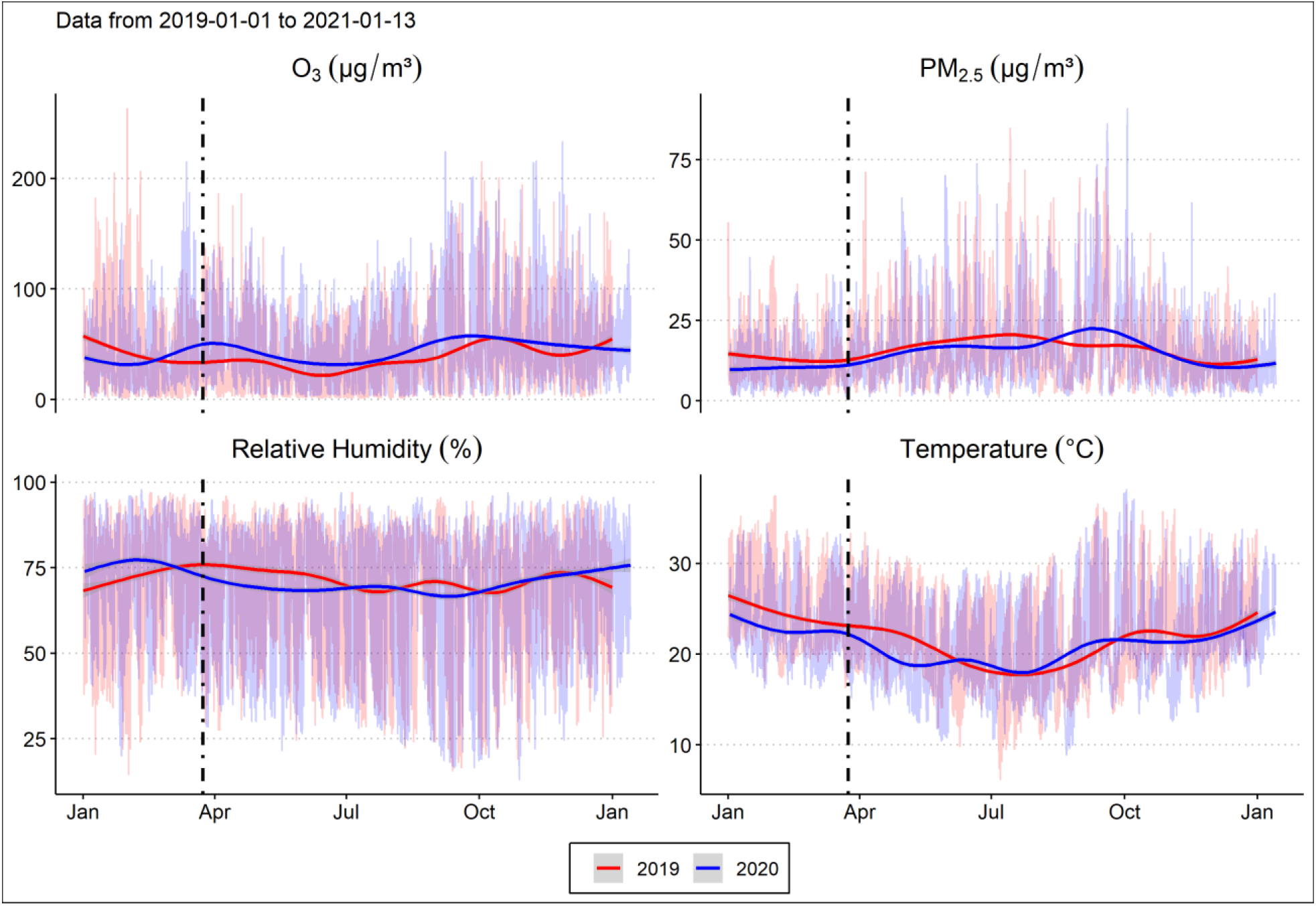
Hourly means of O_3_ (μg/m^3^), PM_2.5_ (μg/m^3^) and Air Temperature (°C) between January 1^st^ 2020 and January 4^th^, 2021 for the city of São Paulo, Brazil (CETESB 2021). The means considered the parameters from the stations Congonhas, Cid.Universitária-USP-Ipen, Santana, Ibirapuera, Mooca, Pinheiros and Parque D. Pedro II. The red and blue lines are the automatic LOESS regression made with ggplot2 and R (R Core Team 2021; Wickham 2016).

### Association between mobility and COVID-19

The statistical analyses consisted of the application of the generalized additive model (GAM) (Hastie and Tibshirani 1990). One of the most common applications of this framework consists of a semi-parametric model in environmental studies to understand the associations between air pollution and health outcomes by controlling other factors such as meteorology (Dominici et al. 2004; Peng and Dominici 2008). Recently, a study has shown that air pollution can increase up to 15% of COVID-19 mortality and worldwide, 27% in East Asia, 19% in Europe and 17% in North America (Pozzer et al. 2020). It has been shown that health effects of air pollution is related to previous days’ exposure (Abrutzky et al., 2013; Carracedo-Martínez et al., 2010; Leitte et al., 2009). In this study we need to consider the incubation period for COVID-19, that is the period of time between the exposure to SARS-CoV-2 and the symptom onset. Furthermore, in this study we want to characterize how the exposure, measured by the mobility indices, is associated with COVID-19. It has been reported that the incubation period for COVID-19 is 5.1 days (95% CI, 4.5 to 5.8 days (Kraemer et al. 2020; Lauer et al. 2020), Lai et al (2020) found between 2 and 14 days with a mean of 6.4 days another (Lai et al. 2020). Therefore, we calculated moving averages between 4 and 21 days of mobility and environmental factors, and study possible associations with COVID-19. We used thin plate splines for accounting confounding factors of PM_2.5_, O_3_, temperature, relative humidity, day of the week and time, including interactions between the variables with quasi-poisson and negative binomial distributions to capture over-dispersion (Wood 2017; Zeileis et al. 2008). For instance, tropospheric O_3_ is a secondary pollutant generated by reactions between NO_X_, Volatile Organic Compounds and solar radiation (Jacob 1999), and as the diurnal cycle of temperature follows solar cycle, we would expect statistical interactions between O_3_ and temperature. To identify associations between mobility and COVID-19, by controlling confounding factors, we used the general equation 1. We performed a detailed sensitivity analyses between the variables, shown supplementary material S1 expanding the equation 1 into 18 configurations.

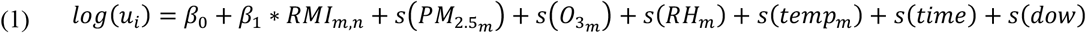

Where log(u) is the log-transform of the daily cases and deaths of COVID-19 with a quasi-poisson or negative binomial distribution *i, β*_0_ is the intercept *β*_1_ is the coefficient that represents the association of *RMI*_*m,n*_ moving average m using mobility data n from Google or SIMI-SP on cases, *s* the thin plate, and *temp* temperature, RH relative humidity, PM_2.5_ and O_3_ atmospheric pollutants, *time* represents each day to account unobserved factors, s(dow) is the cubic spline function with dimension of 7 to account for each day of the week. We used thin-plate splines to avoid knot placement and, therefore, avoid overfitting (Wood 2003). The predicted number of COVID-19 cases is then *exp*(β_0_ + β_1_ ∗ *RMI*_*l*_), which is conditionally to the other predictors. Although more pollutants are reported by São Paulo (QUALAR) air quality stations, e.g., NO_2_, CO, and PM_10_, we limited the model to PM_2.5_ and O_3_ to avoid multicollinearity between PM_2.5_ and these other species. As the objective of this study estimate the associations between daily residential mobility index (RMI), air pollution, and meteorology, and daily cases and deaths for COVID-19 in São Paulo, Brazil, the data were filtered starting on March 24^th^, 2020. In order to apply GAM, we used the R programming language and the library mgcv (v1.8.31) (R Core Team 2021; Wood 2017). One limitation of our method is the limited sample, consisting of 292 days between March 27^th^, 2020 and December 12^nd^, 2020. Nevertheless, we performed a comprehensive sensitivity analysis, as we evaluated 18 equations with 4-21 moving average lag periods, and compared RMI data from Google and SIMI-SP with quasi-poisson and negative binomial distributions, this resulted in 1296 regressions. To ensure consistency, we repeated the analyses with data only until November 2020 finding similar results.

### 2.1. Association between air pollution and COVID-19

We also investigated the effects of air pollution on COVID-19. In this case, we are interested in evaluating the effect of specific level of air pollution present on the same or previous specific days, rather than the moving average of air pollution. The use of lag models is recommended to identify associations with air pollution and health outcomes (Gasparrini 2011; Peng and Dominici 2008). Then, we used lags to account for the delayed effect of air pollution on COVID-19 with a quasi-poisson and negative binomial distributions (Wood 2017; Zeileis et al. 2008). We used single-lag generalized additive models and different configuration for confounding variables with thin plate splines. To identify associations between air pollution and COVID-19, by controlling confounding factors, we used the general equation 2. A sensitivity analyses is available on supplementary material S2, expanding the equation 2 into 8 configurations.

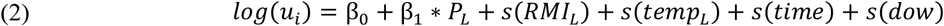

Where P represents the air pollutant concentrations of 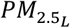 or 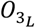, *temp*_*L*_, *RH*_*L*_, and *RMI*_*L*_ are the delayed environmental quantities at different lags l. The methodology is similar as we presented to calculate the effect of the RMI index, but in this case, we controlled all the variables except air pollution. This is useful to compare with other studies and to see the importance of exposure to air pollution and its effects on COVID-19 cases. Then, we calculated the relative risks of new cases by the increment of 10 µg·m^-3^air pollution with the expression exp(β_1_ ∗ *p*ollutant).

## 3. Results

### 3.1. Mobility and COVID-19 cases and deaths

The associations between mobility indices RMI SIMI-SP and Google, parameter β_1_ on equation 1, are shown on Figure 4. The x-axis represents the model configurations from equations 1-20 and the facet labels 4-21 the delayed effect of exposure as moving average. RMI Google was not significantly associated with COVID-19 in any model. This can be explained by the low correlation between RMI Google with Cases -0.19 and with Deaths 0.1 (not significant), as shown on Table 1 on supplementary material. On the contrary, we did find statistical association between RMI SIMI-SP and COVID-19. It is evident increasing the mobility, that is staying out-of-home which results in lower RMI, have a delayed effect increasing COVID-19 cases after four to nine days of exposure. It has been reported that the incubation period for COVID-19 is between 4.5-6.4 days (Kraemer et al. 2020; Lai et al. 2020; Lauer et al. 2020). We believe that the appearance of cases after more days of exposure than the number reported in other studies, is due to the fact that the Reverse transcription polymerase chain reaction (RT-PCR) tests to detect virus takes between 3 days and one week. This explanation also applies to the expected cases after seven, eight, nine and 18 days of exposure. Regarding the deaths, we found associations after 18-21 days of exposure, which makes sense because more time is needed between exposure, severe disease and death outcome. We also found increased deaths after 4 days of exposure, but the magnitude is also lower, near to zero. Furthermore, there is a trend after 14 days of exposure decreasing the β_1_, in which most of models signalized the association with mobility and COVID-19 death. These associations were found with negative binomial and quasi-poisson regressions, and the reader can reproduce these results following the instructions available in this public repository https://gitlab.com/ibarraespinosa/covid191.

**Figure 4.**
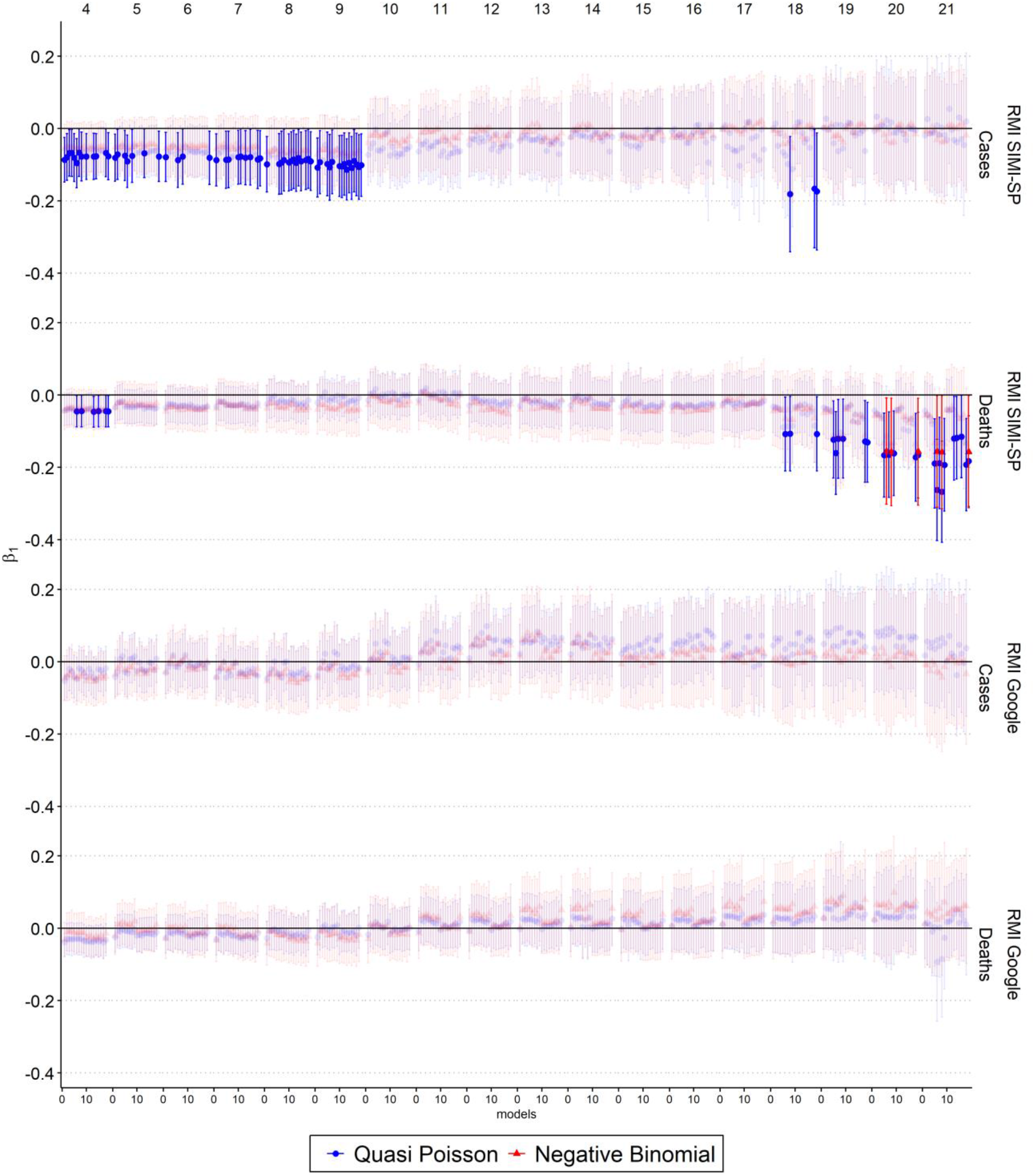
Coefficient of association between RMI SIMI-SP (%) and Google (%) and COVID-19 cases and deaths with a moving average of four to 21 days of delayed exposure with quasi-poisson and negative binomial distribution under different model configuration. β_1_ non-significant associations (p-value > 0.05) are semi-transparent.

After analyzing the coefficients of association RMI and COVID-19, we calculated the expected outcomes with different moving average periods. Then, we grouped the results to obtain one curve exposure-response for cases and deaths, as shown on Figure 5. They grey points represents the expected cases and deaths under different exposure levels, expressed as RMI SIMI-SP and the red curves a smooth LOESS regression with 95% confidence interval for all associations. Our results shows that when people stayed home, COVID-19 cases and deaths decrease. Likewise, with less RMI or increased outdoor activities, there more cases and deaths for COVID-19. The mean RMI SIMI-SP during pre-quarantine (before March 15) was 31.10% and just after the quarantine (between March 27^th^ and April 15^th^, 2020) was 54.77%, increasing the isolation. Then, the association between RMI and cases was assessed with the median RMI SIMI-SP of 45.28% post-quarantine, presented as the second vertical black line in Figure 5. Under this RMI we would expect 1,212 cases (95% CI: 1,189 to 1,235) and 44 deaths (95% CI: 40 to 47). We applied the expected outcomes for several RMI values to evaluate the resulting cases for RMI extremes. For example, under the first quantile of RMI SIMI-SP, first vertical black line, that is 41.38%, it would result in 1,757 cases (95% CI: 1,734 to 1,780) and 80 deaths (95% CI: 77 to 84) and under the third quantile of 48.87%, third vertical black line, it would result in 846 cases (95% CI: 823 to 869) and 25 deaths (95% CI: 22 to 29). Analyzing the extreme values shows that, if the RMI SIMI-SP were 37.82% would result in 2,311 cases (95% CI: 2,285 to 2,338) and 127 deaths (95% CI: 122 to 131) and with RMI SIMI-SP of 59.25%, 351 cases (95% CI: 325 to 378) and 8 deaths (95% CI: 4 to 13). We added a repeated histogram of RMI SIMI-SP on the top of Figure 5, with the RMI quantiles. Therefore, avoiding unnecessary outdoor activities and staying at home would result in a reduction in expected cases and deaths.

**Figure 5.**
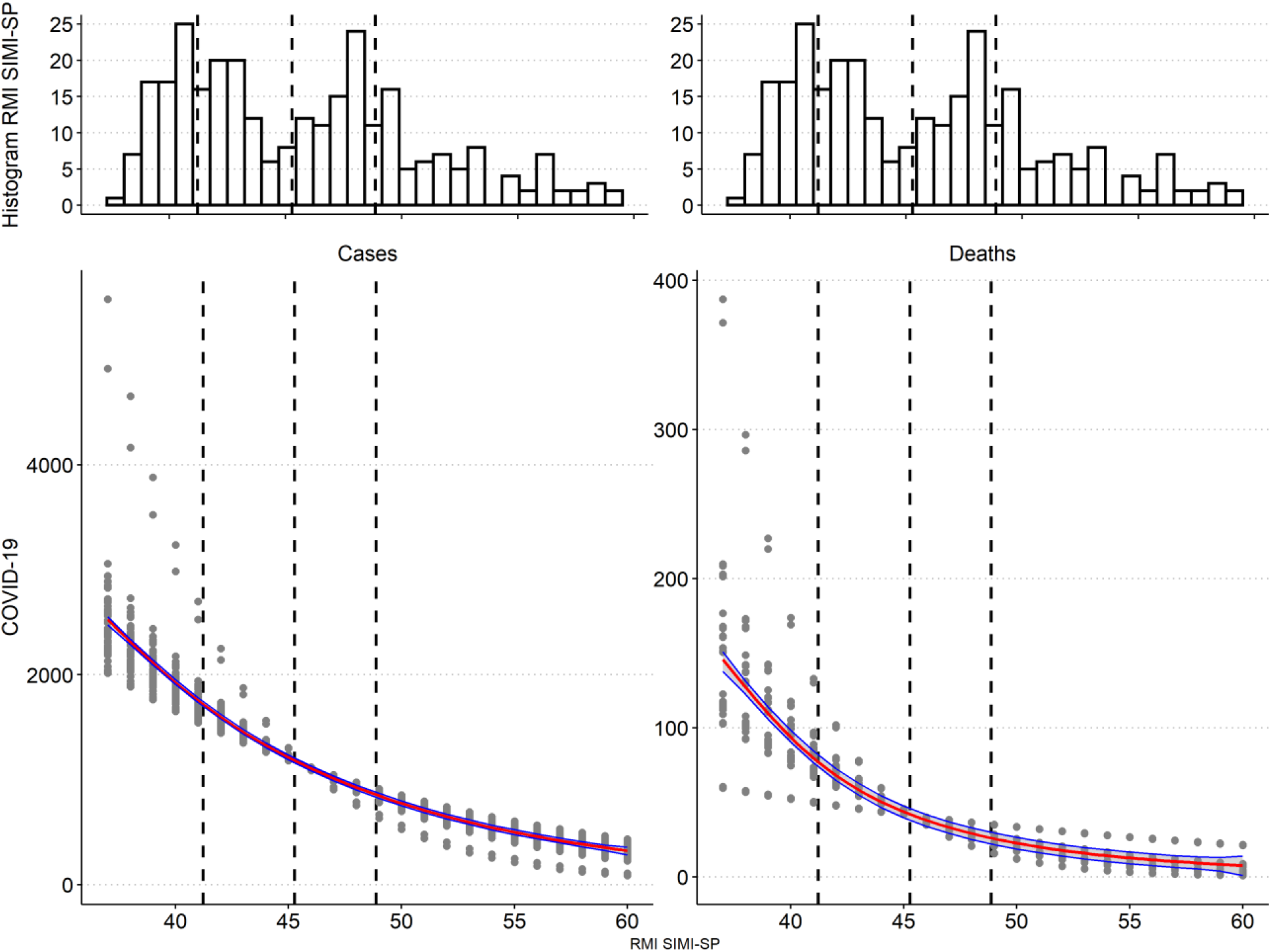
Association between COVID-19 cases and deaths and the percentage of RMI SIMI-SP (%). The grey points show the association between COVID-19 and RMI after different periods of exposure. The red line represents the expected cases and deaths and blue lines upper and lower confidence intervals 95%. The vertical black lines show the first quantile, median and third quantile of RMI SIMI-SP. The histograms of RMI SIMI-SP are repeated.

### 3.2. Air pollution and COVID-19 cases

We assessed the association between COVID-19 and increment of 10 μg·m^-3^ of PM_2.5_ and O_3_. Figure 6 shows the relative risks of COVID-19 cases and deaths after 1-21 days of exposure. We found that PM_2.5_ and O_3_ have positive relative risks for cases and deaths with both distributions. Specifically, O_3_ increments cases after four and 13 days of exposure, and deaths after two, four, 19 and 20 days of exposure, and PM_2.5_ poses positive relative risks after two, 10 and 13 days of exposure for cases and 17 days of exposure for deaths. Nevertheless, there are some relative risks below 1, which would provide protective factors for O_3_ and PM_2.5_ after 17 days for cases where more research is needed. The order of magnitude of relative risks within lagged group is very similar with different models. Therefore, we averaged the relative risks within groups resulting that an increment of 10 μg·m^-3^ of O_3_ produce cases-relative risks of 1.066 (95% CI: 1.005 to 1.131) and 1.084 (95% CI: 1.007 to 1.168) after four and 13 days of exposure, respectively. In the case of deaths, the O_3_-related relative risks are 1.067 (95% CI: 1.008 to 1.129), 1.050 (95% CI: 1.003 to 1.100), 1.070 (95% CI: 1.012 to 1.131) and 1.066 (95% CI: 1.001 to 1.135) after two, four, 19 and 20 days of exposure, respectively. Likewise, an increment of 10 μg·m^-3^ of PM_2.5_ produce cases-relative risks 1.151 (95% CI: 1.048 to 1.264), 1.113 (95% CI: 1.002 to 1.236) and 1.157(95% CI: 1.012 to 1.323) after three, 10 and 14 days of exposure and the risk for death is 1.086 (95% CI: 1.008 to 1.170) after 17 days. Based on these results, air pollution significantly increases COVID-19 cases and deaths. The mean relative risks for cases are 1.140 for PM_2.5_ and 1.075 for O_3_, meaning that PM_2.5_ increments 1.06 times more COVID-19 cases than O_3_. In the case of deaths, the relative risk for O_3_ is 1.063 and for PM_2.5_ 1.086.

**Figure 6.**
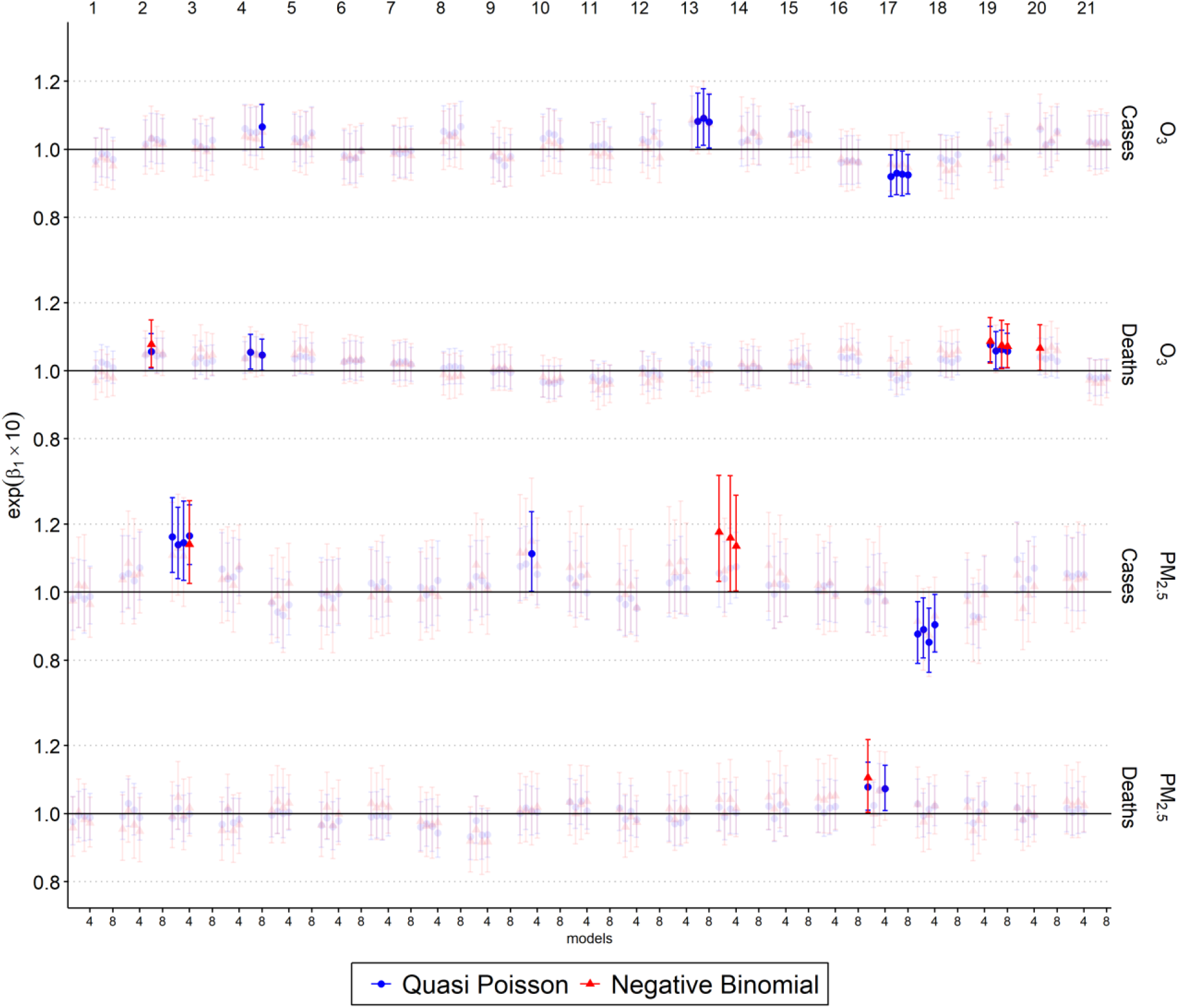
The relative risks of COVID-19 cases and deaths due to 10 μg·m-^3^ of PM_2.5_ and O_3_ after 1-21 days of exposure with single-lag models. β_1_ non-significant associations (p-value > 0.05) are semi-transparent.

## 4. Discussion

In this study, we used a semiparametric Generalized Additive Model (GAM) to explore possible associations between RMI, air pollutants, and COVID-19 cases and deaths in São Paulo, Brazil, for March 27^th^, 2020 through December 30^th^, 2020. We controlled environmental factors such as air temperature, relative humidity, and air pollutant concentrations (PM_2.5_ and O_3_) with thin splines. Arguably, models like the one used here are more commonly applied to much larger datasets. However, we present findings and caveats, to provide early evidence on the transmission of COVID-19, and as part of efforts to highlight the potential value of the recently developed mobility indices. We found statistical associations between RMI and COVID-19 cases and a lower RMI (i.e., the increase of residents staying-at-home) increase COVID-19 cases. Likewise, increased isolation decreases COVID-19 cases. The median RMI after quarantine started was 45.28%, which represents most of the period of study. Under this RMI, we would expect 1,212 cases (95% CI: 1,189 to 1,235) and 44 deaths (95% CI: 40 to 47). São Paulo’s COVID-19 median values are 1,214 and 46 for cases and deaths, which means that our predictions align with our observations and previous literature regarding COVID-19 cases (Martins et al. 2020). We analyzed RMI values to provide policymakers with several options to mitigate the number of COVID-19 cases and deaths and support public health system. Then, if the isolation is increased with the RMI SIMI-SP index from 45.28% to 50.00%, we would expect 774 (95% CI: 751 to 797) cases and 23 (95% CI: 19 to 26) deaths, representing a reduction 438 cases and 21 deaths, a third of the cases and half of the deaths. Therefore, a policymaker can use this information and define RMI targets based on the capacity of their health system.

We evaluated the effect of moving average air pollution on COVID-19 cases and deaths and we found strong associations. The average of the significant relative risk over the 21 days of delayed exposure is 1.140 (95% CI: 1.021 to 1.274) for cases and 1.086 (95% CI: 1.008 to 1.170) for deaths due to an increment of 10 μg·m^-3^ of PM_2.5,_ and 1.075 (95% CI: 1.006 to 1.150) for cases and 1.086 (95% CI: 1.008 to 1.170) for deaths due to an increment of 10 μg·m^-3^ of O_3._ A global study about the association between air pollution and death-risk for COVID-19 found that in South America the attributable fraction (AF) of COVID-19 mortality due PM_2.5_, calculated as 1-1/RR, is approximately 15% in São Paulo (Pozzer et al. 2020), while the AF due the increment of PM_2.5_ in Sao Paulo is 12.28. Another study in China showed that 10 μg·m^-3^ of O_3_ results in relative risk of 1.047 (Zhu et al. 2020a), which is slightly lower than our results of 1.075 presented in this study. Zhang et al (2021) found a country-average relative risk for PM_2.5_ of 1.06 (95% CI: 1.03 to 1.08), while relative risk found in Northeast and Southwest China oscillate around 1.2, similar to our result 1.140 and another Chinese study found a relative risk of relative risk of 1.18 for PM_2.5_ (Zhu et al. 2020b).

As this study is based on 292 days of data, future research should consider the potential effects of a more extended period to study the effect of air pollution on COVID-19 related cases and deaths. Recent social distancing and quarantines have been introduced on unprecedented scales, made necessary by the high transmissivity and severity of COVID-19, and the lack of effective vaccines or testing programs (Cohen and Kupferschmidt 2020). This strongly suggests that mobility indices can be used to study infectious disease transmission and assess the effectiveness of large-scale isolation and quarantine style management activities. Therefore, policymakers can use the new mobility dataset to enforce efforts to implement more effective social distancing and quarantine-based management strategies for COVID-19 in the other states of Brazil.

The use of mask could expand these types of studies, to answer the question if increased mobility using appropriate masks would increases COVID-19. Unfortunately, to the best of our knowledge, there are no time-series available about use of mask in Brazil. Also, the first vaccine applied in Brazil was on January 17, then our study did not include the effects of vaccination (SP 2021). Then, future studies which seek to study the effect of exposure using mobility indices on COVID-19 would need to control newer variables such as, use of mask and number of COVID-19 vaccines applied.

## Conclusion

Spatial distancing was proven to be a determining factor to control COVID-19 cases and deaths. RMI is also significantly associated with COVID-19 cases and deaths. Increased isolation decreases COVID-19 cases while increased mobility is related to a higher number of COVID-19 cases and deaths. Our predictions align with mean observations of COVID-19 cases. Air pollutant models revealed that an increment of 10 μg·m^-3^ of PM_2.5_ and O_3_ produces a relative risk of 1.140 (95% CI: 1.021 to 1.274) for cases and of 1.086 (95% CI: 1.008 to 1.170), and 1.075 (95% CI: 1.006 to 1.150) for cases and 1.063 (95% CI: 1.006 to 1.124) for deaths, respectively.

## Data Availability

All the data used in this manuscript and the scripts to reproduce the results and figures are available at https://gitlab.com/ibarraespinosa/covid191

## Acknowledgments

The authors would like to thank the São Paulo’s Environmental Agency (CETESB), for making possible the use of air quality measurements through its QUALAR platform. The authors would like to thank Lena Goodwin for the support.

## Author contributions

Conceptualization, S.I.E. and E.F.; Methodology, S.I.E, E,F., F.D., K.R., A.R.; Investigation, S.I.E, E.F., K.R., F.D. and A.R.; Writing – Original Draft, S.I.E., K.R. and E.D.; Writing – Review & Editing, S.I.E., K.R., E.F., F.D. and A.R.; Funding Acquisition, E.F.; Supervision, E.F. and F.D;

## Data and materials availability

All the data and code used in this study is available at the only repository https://gitlab.com/ibarraespinosa/covid191.

## Footnotes

1 Climatology between 1981-2010 https://portal.inmet.gov.br/

2 https://www.google.com/covid19/mobility/

