## SupplementaryMaterial for "Negative-Binomial and Quasi-Poisson regressions between COVID-19, mobility and environment in São Paulo, Brazil"

S1. Sensitivity analyses to identify associations between mobility and COVID-19

In order to estimate associations between mobility and COVID-19, a comprehensive sensitivity analyses were performed, by comparing poisson and negative binomial models with a combination adjusted by covariates and their interactions, as shown on equations 1-18. All models included a time variable for each day and also, day of the week. The covariates includes temperature, relative humidity, ozone (O_3_), particulate matter with aerodynamical diameter lesser than 2.5 microns (PM_2.5_) and their interactions via tensor splines(Wood 2006). In addition, we compared mobility trends data from Google based on smart-phones equipped with Global Position Systems (GPS) and data from local cellphone companies, detected by the nearest cellphone antenna, totalizing 1296 regressions. All the code, inputs and outputs of the analyses is found at <https://gitlab.com/ibarraespinosa/covid191> for reproducibility and replicability purposes. The associations were reported as the beta parameter on equations 1-18 and then, the expected cases as the sum of the exponentiated parameter estimates from the models with 95% confidence intervals. All analyses was made using R with the mgcv package (Wood 2017).

(1)

$$log\left( u_{i} \right)=\beta_{0}+\beta_{1}*RMI_{m,n}+s\left( time \right)+s\left( dow \right)$$

(2)

$$log\left( u_{i} \right)=\beta_{0}+\beta_{1}*RMI_{m,n}+s\left( temp_{m} \right)+s\left( time \right)+s\left( dow \right)$$

(3)

$$log\left( u_{i} \right)=\beta_{0}+\beta_{1}*RMI_{m,n}+s\left( temp_{m} \right)+s\left( PM_{{2.5}_{m}} \right)+s\left( time \right)+s\left( dow \right)$$

(4)

$$log\left( u_{i} \right)=\beta_{0}+\beta_{1}*RMI_{m,n}+te\left( temp_{m},PM_{{2.5}_{m}} \right)+s\left( time \right)+s\left( dow \right)$$

(5)

$$log\left( u_{i} \right)=\beta_{0}+\beta_{1}*RMI_{m,n}+s\left( temp_{m} \right)+s\left( O_{3_{m}} \right)+s\left( time \right)+s\left( dow \right)$$

(6)

$$log\left( u_{i} \right)=\beta_{0}+\beta_{1}*RMI_{m,n}+te\left( temp_{m},O_{3_{m}} \right)+s\left( time \right)+s\left( dow \right)$$

(7)

$$log\left( u_{i} \right)=\beta_{0}+\beta_{1}*RMI_{m,n}+s\left( temp_{m} \right)+s\left( PM_{{2.5}_{m}} \right)+s\left( O_{3_{m}} \right)+s\left( time \right)+s\left( dow \right)$$

(8)

$$log\left( u_{i} \right)=\beta_{0}+\beta_{1}*RMI_{m,n}+te\left( temp_{m},O_{3_{m}} \right)+s\left( PM_{{2.5}_{m}} \right)+s\left( time \right)+s\left( dow \right)$$

(9)

$$log\left( u_{i} \right)=\beta_{0}+\beta_{1}*RMI_{m,n}+te\left( temp_{m},PM_{{2.5}_{m}} \right)+s\left( O_{3_{m}} \right)+s\left( time \right)+s\left( dow \right)$$

(10)

$$log\left( u_{i} \right)=\beta_{0}+\beta_{1}*RMI_{m,n}+s\left( RH_{m} \right)+s\left( time \right)+s\left( dow \right)$$

(11)

$$log\left( u_{i} \right)=\beta_{0}+\beta_{1}*RMI_{m,n}+s\left( RH_{m} \right)+s\left( PM_{{2.5}_{m}} \right)+s\left( time \right)+s\left( dow \right)$$

(12)

$$log\left( u_{i} \right)=\beta_{0}+\beta_{1}*RMI_{m,n}+te\left( RH_{m},PM_{{2.5}_{m}} \right)+s\left( time \right)+s\left( dow \right)$$

(13)

$$log\left( u_{i} \right)=\beta_{0}+\beta_{1}*RMI_{m,n}+s\left( RH_{m} \right)+s\left( O_{3_{m}} \right)+s\left( time \right)+s\left( dow \right)$$

(14)

$$log\left( u_{i} \right)=\beta_{0}+\beta_{1}*RMI_{m,n}+te\left( RH_{m},O_{3_{m}} \right)+s\left( time \right)+s\left( dow \right)$$

(15)

$$log\left( u_{i} \right)=\beta_{0}+\beta_{1}*RMI_{m,n}+s\left( RH_{m} \right)+s\left( PM_{{2.5}_{m}} \right)+s\left( O_{3_{m}} \right)+s\left( time \right)+s\left( dow \right)$$

(16)

$$log\left( u_{i} \right)=\beta_{0}+\beta_{1}*RMI_{m,n}+te\left( RH_{m},O_{3_{m}} \right)+s\left( PM_{{2.5}_{m}} \right)+s\left( time \right)+s\left( dow \right)$$

(17)

$$log\left( u_{i} \right)=\beta_{0}+\beta_{1}*RMI_{m,n}+te\left( RH_{m},PM_{{2.5}_{m}} \right)+s\left( O_{3_{m}} \right)+s\left( time \right)+s\left( dow \right)$$

(18)

$$log\left( u_{i} \right)=\beta_{0}+\beta_{1}*RMI_{m,n}+s\left( temp_{m} \right)+s\left( RH_{m} \right)+s\left( PM_{{2.5}_{m}} \right)+s\left( O_{3_{m}} \right)+s\left( time \right)+s\left( dow \right)$$

S2. Sensitivity analyses to identify associations between air pollution and COVID-19

The associations between air pollution and COVID-19 were investigated by applying semi-parametric single-lag models. We performed a sensitivity analyses following the equations 19-26., totalizing 672 regressions. All the code, inputs and outputs of the analyses is found at <https://gitlab.com/ibarraespinosa/covid191> for reproducibility and replicability purposes.

(19)

$$log\left( u_{i} \right)=\beta_{0}+\beta_{1}*PM_{{2.5}_{L}}+s\left( RMI_{L} \right)+s\left( temp_{L} \right)+s\left( time \right)+s\left( dow \right)$$

(20)

$$log\left( u_{i} \right)=\beta_{0}+\beta_{1}*PM_{{2.5}_{L}}+s\left( RMI_{L} \right)+s\left( RH_{L} \right)+s\left( time \right)+s\left( dow \right)$$

(21)

$$log\left( u_{i} \right)=\beta_{0}+\beta_{1}*PM_{{2.5}_{L}}+s\left( RMI_{L} \right)+s\left( temp_{L} \right)+s\left( RH_{L} \right)+s\left( time \right)+s\left( dow \right)$$

(22)

$$log\left( u_{i} \right)=\beta_{0}+\beta_{1}*PM_{{2.5}_{L}}+s\left( RMI_{L} \right)+te\left( temp_{L},RH_{L} \right)+s\left( time \right)+s\left( dow \right)$$

(23)

$$log\left( u_{i} \right)=\beta_{0}+\beta_{1}*O_{3_{L}}+s\left( RMI_{L} \right)+s\left( temp_{L} \right)+s\left( time \right)+s\left( dow \right)$$

(24)

$$log\left( u_{i} \right)=\beta_{0}+\beta_{1}*O_{3_{L}}+s\left( RMI_{L} \right)+s\left( RH_{L} \right)+s\left( time \right)+s\left( dow \right)$$

(25)

$$log\left( u_{i} \right)=\beta_{0}+\beta_{1}*O_{3_{L}}+s\left( RMI_{L} \right)+s\left( temp_{L} \right)+s\left( RH_{L} \right)+s\left( time \right)+s\left( dow \right)$$

(26)

$$log\left( u_{i} \right)=\beta_{0}+\beta_{1}*O_{3_{L}}+s\left( RMI_{L} \right)+te\left( temp_{L},RH_{L} \right)+s\left( time \right)+s\left( dow \right)$$

S3. Pearson correlations between the variables for the period 2020-03-27 and 2021-01-12

We firstly analyzed the correlation between the variables. Table 1 shows the correlation indices between mobility, Cases and Deaths of COVID-19, RMI, O_3_, PM_2.5_ and Temperature. Most correlations were significant (p-value < 0.05). RMI Google had low correlation value with Cases -0.19 and was not significant with Deaths, while RMI SIMI-SP has significant correlations with cases -0.46 and Deaths -0.28. O_3_ has negative and significant correlations with Cases -0.18 and Deaths -0.35, while PM_2.5_ has positive significant correlation only with Cases Deaths -0.16. Temperature only negatively correlated only with deaths -0.11 and relative humidity (RH) was not correlated with COVID-19. These results shows that there is a linear relationship with RMI, with less RMI (more stay out of home), more COVID-19 cases and deaths. Also, with higher PM_2.5_ and less Temperature more COVID-19 cases and deaths.

Table 1. Pearson correlations between the variables for the period 2020-03-27 and 2021-01-12.

|  | Cases | Deaths | RMI Google | RMI SIMI-SP | O_3_ | PM_2.5_ | Temperature |
| --- | --- | --- | --- | --- | --- | --- | --- |
| Deaths | 0.73**** |  |  |  |  |  |  |
| RMI Google | -0.19** | 0.1 |  |  |  |  |  |
| RMI SIMI-SP | -0.46**** | -0.28**** | 0.51**** |  |  |  |  |
| O_3_ | -0.18** | -0.35**** | -0.37**** | -0.13* |  |  |  |
| PM_2.5_ | 0.1 | 0.16** | -0.11 | -0.08 | 0.27**** |  |  |
| Temperature | 0 | -0.11 | -0.27**** | -0.23**** | 0.45**** | 0.38**** |  |
| Relative Humidity | 0.01 | -0.04 | 0.06 | 0 | -0.36**** | -0.61**** | -0.38**** |

Note: "****" is p.value < .0001, "***" is p.value < .001, "**" is p.value < .01 and "*" is p.value < .05 (R Core Team 2020).
